## Supplementary material for "Getting to the Emergency Department in Time: Interviews With Patients and Their Caregivers on the Challenges to Emergency Care Utilization in Rural Uganda - a Grounded Theory Approach": S1 File

### S1 File. Interview Guide

Date:
Time:
Demographic Information:

**Patient:**
First Name:
Gender:
Age:
Parish of Residence:
Occupation:
Highest Level of Education:
None, Primary, Secondary, Tertiary, University
Household Composition – Number of:
- Children (0-12)
- Teenagers (12-18)
- Adults (18-55)
- Elders (55 and up)

**Caregiver:** (brought patient to care, if applicable)
First Name:
Relationship to patient:
Gender:
Age:
Occupation:
Highest Level Education:
None, Primary, Secondary, Tertiary, University

Nyakibale Hospital Emergency Department Visit Information:
Chief Complaint:
Date and Time of Arrival at ED:

Course of Care Seeking (researcher to fill in to ensure all information collected in course of the interview):
Time of onset of illness/injury:
- Location at time of onset:

Time care was first sought outside the home:
- Location at time of decision:

Time of decision to go to ED:
- Location at time of decision:

Interview Questions:
1) When did you/the patient begin to feel unwell, or when did the injury occur?
2) What did you do when you/the patient became unwell, or sustained the injury?
   a) Was care provided at home, or at the scene of an accident?
      i) If yes:
         (1) What care?
         (2) By whom?
3) When did the symptoms become severe enough that someone felt it was necessary to seek healthcare?
4) How was the decision made to seek healthcare?
   a) Who decided that you/the patient needed health care?
   b) Did you (or whomever made the decision to seek care) contact anyone, or ask for advice, prior to seeking medical care?
      i) If Yes:
         (1) Who?
         (2) What advice was given?
      ii) If No:
         (1) Why not?
5) Was care sought elsewhere prior to visiting the Nyakibale Hospital Emergency Department?

i) If yes:
 (1) Where did you go before the ED?
  (a) What treatments were provided?
  (b) What type of transport did you use?
   (i) How much did transport cost?
  (c) How much did the care cost?
 (2) How, and when, did you (or whomever made the decision to seek care) identify the need for emergency medical care?
 (3) Who made the decision to seek medical care at the Nyakibale Hospital Emergency Department?
ii) If no:
 (1) Why not?

6) What barriers to seeking care did you experience for this injury/illness?
 (1) How did you overcome these barriers?
7) Please tell us about your considerations of the following issues before coming to the ED:
 a) Money to pay for care
  i) How much in Ugandan Shillings do you expect to pay for emergency care?
  ii) How did, or will, you obtain money to pay for the care?
   (1) Personal/family savings?
   (2) A loan?
   (3) Selling of resources, such as land, livestock, produce or household things?
  iii) How long did, or will, it take to raise that money?
  iv) Did the fundraising cause a delay in getting care?
   (1) If Yes:
    (a) How long was the delay?
 b) Transport to get to the ED?
  i) What type of transport did you use?
  ii) How did you organize it?
  iii) How much did it cost?
  iv) Did the mobilization of transport cause a delay in getting care?
   (1) If Yes:
    (a) How long was the delay?
 c) Someone to watch children under your care?
  i) How did you organize this?
  ii) Did this cause a delay in getting care?
   (1) If Yes:
    (a) How long was the delay?
 d) Someone to tend your farm, livestock, or business?
  i) How did you organize this?
  ii) Did this cause a delay in getting care?
   (1) If Yes:
    (a) How long was the delay?
 e) Time off from work?
  i) How did you organize this?
  ii) Did this cause a delay in getting care?
   (1) If Yes:
    (a) How long was the delay?
 f) The time of day or night?
  i) Did this cause a delay in getting care?
   (1) If Yes:
    (a) How long was the delay?

g) The weather?
   i) Did this cause a delay in getting care?
      (1) If Yes:
         (a) How long was the delay?
h) What other factors did you consider?
   i) Did this cause a delay in getting care?
      (1) If Yes:
         (a) How long was the delay?

8) What experiences have you had with healthcare, or hospitals, that set your expectations for emergency care?
   a) Have you been to Nyakibale Hospital Emergency Department before?
      i) If yes:
         (1) For what?
         (2) How was your experience?
         (3) What were your expectations for what would occur, and how it would occur, at the ED? In other words, what did you expect from the hospital or ED staff?
      ii) If no:
         (1) How did you get to know about the ED?
         (2) What were your expectations for what would occur, and how it would occur, at the ED? In other words, what did you expect from the hospital or ED staff?
9) What would make it easier for you, and other community members, to make use of the Nyakibale Hospital Emergency Department services?
10) Is there anything else that you would like to tell us?
