## Supplementary material for "Getting to the Emergency Department in Time: Interviews With Patients and Their Caregivers on the Challenges to Emergency Care Utilization in Rural Uganda - a Grounded Theory Approach": S3 File

### S3 File. ED Patients' Chief Complaints

**<u>Medical: 76% (n=38)</u>**

**Neurologic: 18% (n=7)**
Unilateral Weakness
Headache (5)
Dizziness and Headache

**Gastrointestinal/Genital Urinary: 31% (n=12)**
Diarrhea and Vomiting
Epigastric Pain
Abdominal Pain, Swollen Limbs
Abdominal Distention
Lower Abdominal Pain
Severe Abdominal Pain
Abdominal Pain, Vomiting
Abdominal Pain
Lower Abdominal Pain
Lower Abdominal Pain, Headache
Diarrhea and Cough
Painful Scrotum

**Cardiopulmonary: 24% (n=9)**
Severe Chest Pain and Weakness
Chest pain, Headache and Constipation
Productive Cough
Difficulty Breathing
Diarrhea and Difficulty Breathing
Difficulty Breathing, Nausea and Vomiting
Chest pain
Chest pain
Weakness and chest pain

**Other: 26% (n=10)**
Facial Swelling
Severe Epistasis
Fever and Vomiting
Fever and Cough
Fever and Vomiting
Fever
Painful Finger
Neck Swelling
Painful Arm
Fever

**<u>Trauma: 24% (n=12)</u>**

**Burns: 25% (n=3)**

Burns to Face, Chest
Burns to Face, Chest and Bilateral Arms
Burns to Bilateral Feet

**Road Traffic Accidents: 25% (n=3)**

Laceration Ear, Severe Headache, Nausea and Vomiting
Right Arm Pain
Painful Amputated Finger

**Blunt Trauma/Falls: 42% (n=5)**

Epigastric pain Related to Assault
Left Leg Pain Related to Slip and Fall
Chest pain and swollen lips Related to Fall
Painful Shoulder Related to Assault
Painful Leg Related to a Slip and Fall

**Bites: 8% (n=1)**

Painful Hand Related to Dog Bite
