## Supplementary material for "Getting to the Emergency Department in Time: Interviews With Patients and Their Caregivers on the Challenges to Emergency Care Utilization in Rural Uganda - a Grounded Theory Approach": S2 File (Revised)

**S2 File. Code Book with Supplemental Quotes**

### Cultural Factors and Limited Knowledge of Emergency Signs and Initial Actions to Take

**Decision to Seek Care**

**Head of family, educated or medical, or financial resources to fund care leads to decision making**

“When I told the father of the patient that I was going to take the matter to the police or leave the patient at his home then he called the BODABODA cyclist who brought us to the emergency department” (20-29yo parent seeking care for <5yo child)

“(My sister who is a nurse) is the one who called our brother (to tell him) that the patient’s symptoms had become severe and then our brother later called me to bring the patient here (to) Nyakibale hospital… My sisters and my brothers are the ones to arrange and pay for the care because they educated and have some money” (40-49yo child seeking care for an 80-89yo parent)

**Wait to be advised/seek opinions**

“The neighbor saw the patient and said you go to the hospital. They organized for the BODABODA that brought me here for a debt. They are to pay.” (20-29yo parent seeking care for 5-9yo child)

“I think failure to make a decision to seek medical care delayed me, because one was saying this and another saying that.” (40-49yo patient seeking care)

“Of course I knew the disease and I had the medical papers from the previous medical care Centre’s. Therefore, I never needed to ask for any advice” (40-49yo parent seeking care for 20-29-yo child)

**Difficulty Identifying Illness/Injury Amenable to ED Care**

**Perception of traditional illness**

“There are other diseases that attacked her that come from their family, the family of my husband. We first thought that the patient was suffering from traditional disease and therefore we had to first try treating with traditional herbs and then (go) to clinic.” (20-29yo parent seeking care for <5yo child)

“Some of people advised me that such kind of illness would be well managed by traditional herbs. I said “let me first seek medical care from health centers and if it fails then (that is) when I will try traditional herbs”. Actually, previously I had tried traditional herbs because I had (sought) medical care from here (Karoli Lwanga Hospital), and also from the clinics, but it never helped the patient and it was then that I used those traditional herbs.” (30-39yo parent seeking care for 5-9yo child)

“My husband first considered whether the patient’s illness was a traditional disease so that I can get her a traditional healer because I have sought medical care all over the health centers. The patient was put into scan and given other treatment but it never helped. But, still I insisted that it wasn’t a traditional diseases and therefore we needed to come here.” (30-39yo parent seeking care for 10-19yo child)

“The clinical personnel advised (the patient’s mother) that the illness was not a modern disease and therefore she needed to take the patient to the church for prayers or else traditional healers. Actually that was why the mother of the patient was insisting that she knew the illness, but I disagreed with her saying that that clinical personnel doesn’t have any medical equipment to diagnose a traditional illness.” (20-29yo seeking care for a 20-29-yo relative)

“When I reached at the village, the patient couldn’t recognize me as he was always looking down and when they saw that they thought it was Satan that disturbed him…I just took him for prayers for two days and after realizing that he was not getting any improvement then I decided to bring him (to the ED).” (20-29yo seeking care for a 20-29-yo relative)

“I had talked to the patient in the morning about the idea of bringing him here but he was adamant saying that “You see I have taken Tayebwa (a local herbal remedy), the nurses and doctors would be hard on me. Let it first come out.” I said “What if you don’t pass it out, what will you do?” That’s when I called the children and told them that your father is becoming worse, they called him and told him that “You need to be taken to Nyakibale now.” (70-79yo seeking care for 70-79yo spouse)

**Chronic Illness**

“I think money didn’t delay us but the problem was that we thought the patient would become well since we were treating the right disease with the right drugs and actually this gave us hope that the patient never needed any other medical care” (60-69yo parent seeking care for her 30-39yo child)

“When I called, (some) said, wait we are still looking for money, (but) others say no, AIDs can’t be cured let her die….(laughs)…yes” (30-39yo seeking care for 30-39yo relative)

“My husband had lost hope in trying to seek medical care for her because he was complaining that he started seeking medical care for the patient since she was two years up to now he had not seen any improvement. Therefore, he was saying that there (would be) no change with coming here (to) Nyakibale hospital for medical care again” (20-29yo parent seeking care for 10-19yo child)

**Decision making after patient becomes severe**

“On Saturday night around 2:00am the symptoms became very severe. I couldn’t breathe well, I didn’t sleep and I was like “I am going to die here”… I said “let me call a bodaboda (motorcycle taxi) to bring me so that I die from the hospital.” It is from then that I made a decision to come to Nyakibale (Hospital).” (30-39yo patient)

**Decide after other care fails, or they are referred**

“After we had sought medical care from the other health facilities, like government health facility, and (they) failed then we decided to come here.” (40-49yo parent seeking care for 10-18yo child)

**Use of Local Facilities for Stabilization and Advice Despite Perception of Inadequate Services**

“Actually money for the care here is always biggest problem and it scares away many people (away from) coming here for care. Of course people are different, some can afford and some can’t afford… But, medical care here is extremely unaffordable and that’s why many people don’t come here.” (40-49yo seeking care for 20-29yo child)

“The patient had refused to be brought here because he was complaining that he never had money for medical care and then we had to bring him here by force.” (50-59yo child seeking care for 90-99yo parent)

“The biggest problem was the money problem because I am the only one taking care of the family.” (30-39yo parent seeking care for 5-9yo child)

“Of course with money you can do anything at the right time and without it you can’t because you fear doing something…it delayed us because you fear if I am taken to the hospital and I also have to pay for school fees” (20-29yo child seeking care for 50-59yo parent)

**Treatment before payment**

“Right now we are still soliciting for money. We thought about her life first, before thinking about money for medical care here…We were like “we will look for money later but let us first take her to the hospital”.” (20-29yo child seeking care for 60-69yo parent)

“I can’t say that I have overcome the barriers of money yet, because I can’t sit here and say I have money to pay…I don’t know how I will still get the money after the hospital has billed us…I still have to look for the money by calling my friends” (30-39yo seeking care for 40-49yo spouse)

**Limited first aid or home care**

“We were only providing good diet to the patient like fish because the patient couldn’t eat sweet potatoes…. (we) boiled water with traditional herbs and massage the joints but it never helped.” (50-50yo seeking care for 80yo parent)

“When the patient fell into the fire and got burnt, he had been left home with his brother, who quickly splashed cold water on the burns trying to help the patient, but the skin just got off.” (40-49yo parent seeking care for <5yo child)

**Rush to local clinics**

“When the patient felt unwell, we took the patient to the clinic. (We) were admitted from Wednesday up to Friday when the patient was discharged. Then on Sunday the patient got severe again and I took the patient back to the clinic…but the symptoms never improved and then we decided to come here.” (20-29-yo parent seeking care for <5yo child)

“At the time the patient got unwell I was not at home…When I came home I quickly rushed her to the clinic but no sooner had she started taking the drugs, the symptoms got severe enough and I decided to bring her here.” (20-29yo parent seeking care for 5-9yo child)

“They (other drivers) didn’t do anything (first aid), they just rushed him to the clinic.” (20-29yo seeking care for 20-29yo spouse)

“The area was remote and therefore no one did first aid, only the traditional healer did traditional incisions and actually also trying to set the born.” (40-49yo patient seeking care)

**Use of Near Facilities**

“It was lack of money, if you don’t have money then you decided to stay home or go to private clinics because you calculate the money for transport to getting here (the ED)….then you decide to go to nearest clinic.” (60-69yo patient seeking care)

“If it weren’t the fundraising for upfront (cost) then we wouldn’t have taken the patient to clinics and government health facility.” (30-39yo parent seeking care for 10-19yo child)

“We took the patient to clinics because we never had enough money to bring him here. So, we felt like let us first seek care at the clinic so that they can give him medication to sustain him as we are arranging to come here. But, still the medication at the clinic never helped the patient and we continued having the wish to come to the emergency department.” (30-39yo seeking care for 40-49yo spouse)

**Use of multiple facilities**

“If you seek medical care from other health care centers and the patient gets well you do not take them back. If patient fails to get well, then you decide to go to different hospital for different medical care and drugs.” (40-49yo parent seeking care for 10-19yo child)

“When the patient got burnt that day we went to the small private clinic the patient was given some injections and admitted for two days. We were later discharged back home but after one week the patient became severe again. This time we decided to take the patient to the bigger private clinic. Reaching there the patient was given medical care and we were later discharged back home after two weeks. Starting with this week, the clinic personnel has been advising us to take the patient back for dressing but we saw that going back to the private clinic was no longer helping us.” (40-49yo parent seeking care for <5yo child)

**Delay in referral**

“The symptoms became severe enough after we had reached at the clinic on Tuesday…We slept there for a night and on the following morning, the clinic nurse said she couldn’t manage the patient. Therefore, she referred us here.” (20-29yo child seeking care for 50-59yo parent)

“At other hospitals they said that she is too weak, she should be brought to (the ED), but others said the money is a lot and you can’t manage (it)… or she will be fine.” (30-39yo seeking care for 30-39yo relative)

“Actually the clinic personnel had earlier told us that if (the patient) continued with serious vomiting, dizziness or severe headache then he would consider referring us for further management. But, the clinic staff member was not ready to refer us (to the ED)…he was still insisting that he had sent someone for good and strong drugs from the pharmacy therefore we were to wait for those good drugs.” (20-29yo seeking care for 20-29yo spouse)

“I took her to Itojo hospital but actually before I took her there, I had to first sought healthcare in many private clinics…I decided to take the patient to Itojo hospital and reaching there, they referred us here…I took the patient back home (for 2 days) so that I could prepare and come here….I wanted to first prepare for what we were to use here like food, and other things.” (20-29yo parent seeking care for <5yo child)

“They gave us few tablets and they also referred us here, but (we) went back home (for 2 months) because of lack of money.” (30-39yo seeking care for 30-39yo relative)

**Inadequate medical services at high cost**

“They were not caring about the child. They would just tell me to put him there she will be well, or say “cover and dress the patient so that she can sweat and get well”. Then I saw that it was not going to help." (20-29yo parent seeking care for <5yo child).

“I had bad experiences with the previous health care Centre’s because I spent too much money…but it did not yield any good results. I also spend a lot of time doing nothing because this delayed us a lot. If we had come earlier then the severity of patient wouldn’t be there.” (40-40yo parent seeking care for <5yo child)

“At the clinics they charge us expensively but the patient gets better for few days and gets sick again in just a short period of time. But, when you seek medical care here at Nyakibale hospital then you patient stays long minus getting sick again.” (20-29yo parent seeking care for <5yo child)

**Lack of explanation of patient’s condition**

“The clinic personnel never gave us any other information regarding whether we needed to take the patient back for further management and also the drugs from the clinic never helped. It from then that I decided to seeking medical care here.” (30-39yo parent seeking care for 5-9yo child)

**No medical attention and services at government facilities, although free**

“I went to the government health Centre after I had sustained the injury. (The nurse) told me that she would like to help me but unfortunately they did not have even a single drug in the health center…I went back home because I never had money. Then on Saturday night when the symptoms became severe that’s when I decided to come here for medical care.” (30-39yo patient seeking care)

### Lack of Resources to Cover the Direct, Indirect, and Opportunity Costs of EC

**Funds for upfront cost of seeking care**

“Money, lack of money is always a problem because you need to organize a car to bring the patient here when you have no money… We have not (obtained) the money for the medical care, but when we got the money for transport we brought the patient here.” (50-59yo child seeking care for 80-89yo parent)

“Sometimes you get sick when you don’t have money and the moment you try calling someone with a vehicle to transport you to the health facility then he says “you first pay me money”.” (40-49yo patient seeking care)

“The big barrier was that whenever I used to tell my husband he would say that “he never had money for the transport” and I would also advise him at least to fundraise money for transport so that I can come with the patient. Also, I told him that if we get money for transport and get to the hospital then we can fundraise money for the medical bill later.” (20-29yo parent seeking care for 10-19 yo child)

“I actually made the decision (to seek ED care) on Friday but unfortunately I never had money, therefore I had to wait and organize money for transport, (delaying us) until Monday.” (40-49yo parent seeking care for 20-29yo child)

“My neighbor had a friend who is a cyclist so he called him and said “the child of my neighbor is badly off, can you help me and drop her at the hospital.” (20-29yo parent seeking care for 5-9-yo child)

“The person who brought us was coming to Rukungiri and he is a village mate. He was coming to buy some things in town and he decided to bring us.” (20-29yo parent seeking care for <5yo child)

“I used BodaBoda (motorcycle taxi) … because of the patient’s condition I always save whenever I get some little money, but still, because I am the only person taking care of the family, it is always difficulty.” (30-39yo parent seeking care for 5-9yo child)

**Family, property, farms and business responsibilities**

“I am the one responsible for my farms. I was harvesting millet and eventually got so sick I couldn’t stand. Actually, this caused a long delay, for four days, because I was thinking about who will take care of my farms. I had even reached the point of refusing to be taken for medical care. I was worried about what I will feed my children after being discharged from the hospital, because I was leaving all my harvests in the farm…This disturbed (me) a lot.” (40-49yo patient seeking care)

“Of course I never needed time off (from work), because when someone gets sick of course you have to leave work and take the patient for medical care. Obviously, you can’t go to work and leave someone sick at home.” (50-59yo child seeking care for 90-99yo parent)

### Inadequate Transportation Options, Especially at Night and in Inclement Weather

**Cost of transport**

“I took a BodaBoda but no sooner had we traveled one kilometer (then) I again developed difficulty in breathing, and therefore I couldn’t move on BODABODA anymore. (Then) they called a car that brought me here.” (60-69yo patient seeking care)

“I have been getting medical care here at nyakibale emergency department and I think its affordable but the problem is when you add the transport costs from village to getting here.” (20-29yo parent seeking care for <5yo child)

**Lack of transport means**

“From home we used a bicycle up to the road, then got a taxi…because its far, you have to wake up at 5 am and go to the road, which is also a problem.” (30-39yo seeking care for 30-39yo relative)

“It is around one mile from our home to the main road and there are few BODABODAS and taxis on that road. Therefore, we had to first walk to reach the main road, where we got a (shared) taxi…It delayed us because if there was car that could reach our home then we would have reached (the ED) very early in the morning” (40-49yo parent seeking care for <5yo child)

“We were coming from far and we had failed to find a Bodaboda, until we got one that transported us (as far as) a trading center. (There) we waited for a (shared) taxi but we couldn’t find one…We decided to use another BODABODA up to here.” (40-49yo child seeking care for 80-89yo parent)

“Where I come from its hard to find easy means of transport, because to get a (shared) taxi to bring you here it has to be in the morning… beyond 8:00am you have to hire a Bodaboda, which is expensive, or wait for the next day.” (20-29yo parent seeking care for <5yo child)

**Night**

“The roads are in very bad condition to travel at night, actually its very risky.” (20-29yo seeking care for 20-29yo spouse)

“It was not easy to drive at night and the roads are not good, with many corners. Therefore, we were risking to drive at high speed because they never wanted to find when the medical workers had closed” (40-49yo patient seeking care)

“Night caused a delay because where we come from is far and the transport is not good and the patient got severe headache at night around midnight, therefore I had to first wait until it was morning.” (30-39yo seeking care for her 40-49yo spouse)

“We did not sleep for the whole night in trying to care for the patient up to morning and actually if it wasn’t night then we would have reached here earlier.” (40-49yo parent seeking care for <5yo child)

“Initially we had decided to bring the patient at night but wasn’t possible because it is not easy to find BODABODA at night, and still, if you happen to get one then he can charge you expensively.” (60-69yo parent seeking care for 30-39yo child)

“You see where we come from, in case someone feels unwell or gets an injury at night it is very hard to get transport means, because you can’t wake up a Bodaboda cyclist to take you for medical care and it is far.” (40-49yo parent seeking care for <5yo child)

“The big problem is money because even if the patient gets severe at night if you have money then you can bring the patient. You see, if you don’t have any generating income factor, or a small business where you can fetch some money, then it is always difficult… Actually, very many people in our village have vehicles but also if it’s night then they can’t accept to be hired to transport patients to the hospital.” (40-49yo parent seeking care for 10-19yo child)

“(We) called the owner of the vehicle…he failed to pick because it was at night and he doesn’t want his vehicle to move at night.” (70-79yo seeking care for 70-79yo spouse)

**Bad Weather**

“If it rains you cannot use a Bodaboda. You can’t even get transport means because roads are always muddy.” (30-39yo parent seeking care for 10-19yo child)

“This time around weather never caused a delay, but previously I was taking the patient to the government health facility and it rained. I had to cancel the program and take the patient the next day.” (20-29yo parent seeking care for 10-19 yo child)
